# Trends over time in the risk of adverse outcomes among patients with SARS-CoV-2 infection

**DOI:** 10.1101/2021.03.08.21253090

**Authors:** George N. Ioannou, Ann M. O’Hare, Kristin Berry, Vincent S Fan, Kristina Crothers, McKenna C. Eastment, Emily Locke, Pamela Green, Javeed A. Shah, Jason A. Dominitz

**Author notes:** **Please direct correspondence to:** George N. Ioannou, BMBCh, MS, Veterans Affairs Puget Sound Healthcare System, 1660 S. Columbian Way, Seattle, WA, 98108, Twitter: @gnioannou. **Funding:** The study was supported in part by the Department of Veterans Affairs, CSR&D grant COVID19-8900-11 to GNI. **Declaration of Personal Interests:** None of the authors has any conflicts of interest to disclose. **Disclaimer:** The contents do not represent the views of the U.S. Department of Veterans Affairs or the US Government. **Authors’ Contributions and Authorship Statement** George Ioannou is the guarantor of this paper. All authors approved the final version of the manuscript. Ioannou: Study concept and design, analysis of data, interpretation of results, drafting of manuscript, critical revision of manuscript, obtaining funding. Locke: Study concept and design. Green: Extraction of data, creation of analytic variables, analysis of data, interpretation of results. Berry: Study concept and design, analysis of data, interpretation of results, drafting of manuscript, critical revision of manuscript. O’Hare: Study concept and design, interpretation of results, critical revision of manuscript. Shah: Study concept and design, critical revision of manuscript. Crothers: Study concept and design, interpretation of results, critical revision of manuscript. Eastment: Study concept and design, interpretation of results, drafting of manuscript, critical revision of manuscript. Fan: Study concept and design, interpretation of results, critical revision of manuscript. Dominitz: Study concept and design, interpretation of results, drafting of manuscript, critical revision of manuscript.

## Abstract

**Objectives:** We aimed to describe trends in the incidence of adverse outcomes among patients who tested positive for SARS-CoV-2 between February and September 2020 within a national healthcare system.

**Setting:** US Veterans Affairs national healthcare system.

**Participants:** Enrollees in the VA healthcare system who tested positive for SARS-CoV-2 between 2/28/2020 and 9/30/2020 (n=55,952).

**Outcomes:** Death, hospitalization, intensive care unit (ICU) admission and mechanical ventilation within 30 days of testing positive.The incidence of these outcomes was examined among patients infected each month and trends were evaluated using an interrupted time-series analysis.

**Results:** Between February and July 2020, during the first wave of the US pandemic, there were marked downward trends in the 30-day incidence of hospitalization (44.2% to 15.8%), ICU admission (20.3% to 5.3%), mechanical ventilation (12.7% to 2.2%), and death (12.5% to 4.4%), with subsequent stabilization between July and September 2020. These trends persisted after adjustment for sociodemographic characteristics, comorbid conditions, and documented symptoms and after additional adjustment for laboratory test results among hospitalized patients, including among subgroups admitted to the ICU and treated with mechanical ventilation. Among hospitalized patients, use of hydroxychloroquine (56.5% to 0%), azithromycin (48.3% to 16.6%) vasopressors (20.6% to 8.7%), and dialysis (11.6% to 3.8%) decreased while use of dexamethasone (3.4% to 53.1%), other corticosteroids (4.9% to 29.0%) and remdesivir (1.7% to 45.4%) increased from February to September.

**Conclusions:** Among patients who tested positive for SARS-CoV-2 in a large national US healthcare system, risk for a range of adverse outcomes decreased markedly between February and July, with subsequent stabilization from July to September. These trends were not explained by changes in measured baseline patient characteristics.

## Introduction

Infection with SARS-CoV-2 results in a broad spectrum of clinical severity ranging from asymptomatic infection to life-threatening illness.^1, 2^ Among individuals who test positive for SARS-CoV-2, there is substantial variation across different populations in rates of hospital admission (from 8% to 80.7%),^3-5^ mechanical ventilation (from 2.3% of the Chinese population to 93.2% of critically ill admitted to New York area hospitals^6-18^), and mortality (from 2.8% of all patients in the US to 10.2-67% of hospitalized and high-risk patients).^3, 7, 8, 12, 17-23^ To date, only a few studies have described trends in these measures over time within populations.^24-26^

Since the first case of SARS-CoV-2 was diagnosed in the US on January 19, 2020, the pandemic has spread across the country from early epicenters in the Pacific Northwest, New York, New Orleans and Detroit. During this time, there have been marked changes in the availability and approach to testing, in population screening strategies, in mitigation strategies both in the community and within health systems, in how symptomatic patients are managed medically and most recently in the availability of vaccination. However, even prior to the availability of a vaccine and as the number deaths due to COVID-19 across the world continued to climb^27^, several studies described improved outcomes among infected patients after the initial months of the pandemic. Improvements in mortality were reported for patients treated in an 8-hospital health system in Houston, Texas between mid-March and mid-July, in a 3-hospital system in New York between March and August, 2020^26^, and in a single hospital in Milano, Italy between late February and mid-May, 2020.^24, 28^ Similar improvements in outcomes over time have also been described for the most severe cases, including among patients admitted to the ICU in England, Wales and Northern Ireland between February and May, 2020 and among patients admitted to the ICU in England from March through June, 2020.^25, 29^ To our knowledge, no prior studies have described temporal trends in the range of severe complications of COVID-19 including hospital admission, intensive care unit (ICU) admission, mechanical ventilation, dialysis and death in a national US health care system and extending beyond the first wave of the pandemic i.e. after June 2020. The Department of Veterans Affairs (VA) healthcare system supports the largest, integrated, comprehensive national healthcare system in the US, providing a unique view of national trends in outcomes among those infected with SARS-CoV-2. We used national data from the VA to measure trends in hospitalization, ICU admission, mechanical ventilation, dialysis, medication use and death among individuals who tested positive for SARS-CoV-2 between February 28 and September 30, 2020, with follow-up extending through November 19, 2020.

## Methods

### Data source and study population

The VA provides care for more than six million Veterans annually and uses a single, national comprehensive electronic healthcare information network. We used data from the VA’s Corporate Data Warehouse (CDW), a relational database of VA enrollees’ electronic health records (EHR), developed by the VA Informatics and Computing Infrastructure (VINCI) to support research and clinical operations, including datasets and analytic variables specifically related to SARS-CoV-2 developed by VINCI as part of the “COVID-19 Shared Data Resource.”^30^.

This study was approved by the Institutional Review Board of the Veterans Affairs Puget Sound Healthcare System, which granted a waiver of informed consent.

### Study Population, definition of SARS-CoV-2 infection and index date

We identified all VA enrollees (n=559,616) with available results of testing for SARS-CoV-2 using approved polymerase chain reaction (PCR) tests from 02/28/2020 (the date of the first SARS-CoV-2 test in the VA system) to 9/30/2020, including 55,952 (10.0%) who tested positive at least once. Final adjudication of SARS-CoV-2 status was performed by the VA National Surveillance Tool, which is intended to be the single, authoritative data source for determination of positive and negative cases within VA. The index date was the earliest date of a documented positive test or, in rare instances, if the patient was already hospitalized prior to testing positive, the date of hospitalization.

### Adverse outcomes

We ascertained the following four outcomes within 30 days of the index date: 1) hospitalization, 2) ICU admission, 3) mechanical ventilation, and 4) all-cause mortality. Deaths that occurred both within and outside the VA are comprehensively captured in CDW through a variety of sources including VA inpatient files, VA Beneficiary Identification and Records Locator System (BIRLS), Social Security Administration (SSA) death files, and the Department of Defense.^31^ However, episodes of mechanical ventilation, ICU admission and hospitalization that occurred outside the VA are captured only if these were paid for by the VA and thus do not include episodes of care covered by Medicare, Medicaid, the Department of Defense or private health insurance. Patients were followed through 11/19/2020 for study outcomes allowing for a minimum follow-up time of 50 days from the most recent index (09/30/21), which was adequate for outcomes to be electronically recorded in CDW.

### Baseline characteristics

Baseline *sociodemographic characteristics*, previously associated with adverse SARS-CoV-2 outcomes,^32^ included age, sex, race, ethnicity, body mass index (BMI), urban vs. rural residence (based on zip codes), and geographic location (divided into 10 US Federal Regions^33^) (**Table 2**).

*Comorbid conditions* were based on ICD-10 codes recorded in VA electronic health records during the 2-year period before the index date.^30^ We used the Deyo modification^34^ of the Charlson Comorbidity Index (CCI)^35^ to estimate overall burden of comorbidity (**Table 2**). We also extracted the following comorbid conditions: diabetes, chronic kidney disease, hypertension, obstructive sleep apnea, and obesity hypoventilation, which were associated with adverse outcomes among SARS-CoV-2 infected Veterans.^32^

We identified *symptoms* potentially associated with SARS-CoV-2 documented on or within 30 days prior to the index date based on a combination of natural language processing to capture relevant documentation in the EHR and searching for relevant ICD-10 codes,^30^ occurring on or within 30 days prior to the index date. In our multivariable models, we used only two symptoms (dyspnea and fever).

Finally, we extracted 5 routine *laboratory blood tests* (albumin, aspartate aminotransferase, creatinine, white blood cell count, neutrophil-to-lymphocyte ratio), which we recently showed to be associated with adverse outcomes in SARS-CoV-2^32^. For each test, we extracted the value closest to the index date, on or within 10 days before the index date, or if absent, within 5 days after the index date (93% of tests were performed within 2 days of the index date).

### Pharmacotherapy, Noninvasive Ventilation and Dialysis

We identified medications prescribed for SARS-CoV-2 on or within 60 days after the index date. We also report use of non-invasive ventilation (continuous (CPAP) or bilevel (BiPAP) positive airway pressure) and dialysis during the same time frame. Use of high-flow oxygen supplementation could not be reliably captured in available data sources for this project.

### Statistical Analysis

We divided the observation period into monthly intervals based on the index date. Using the Kaplan-Meier method, we calculated for the persons infected each month the 30-day cumulative incidence of hospitalization, ICU admission, mechanical ventilation, and mortality from the index date. Patients who did not experience the relevant outcome of interest were censored at the time of death or after 30 days of follow-up. We used Cox proportional hazards regression to compare the risk of each outcome among patients infected during each subsequent month relative to the earliest time period after adjusting for the following baseline characteristics, which we found in a prior study to be associated with adverse outcomes among SARS-CoV-2-infected VA enrollees^32^: age, sex, race, ethnicity, BMI, regional COVID-19 disease burden, CCI, diabetes, chronic kidney disease, hypertension, obstructive sleep apnea, obesity hypoventilation, fever, and dyspnea. In analyses limited to hospitalized patients, we additionally adjusted for the 5 laboratory tests listed above that have also been associated with adverse outcomes^32^. Cox proportional hazards analyses were stratified by the VA medical center where patients were tested for SARS-CoV-2. The aim of these multivariable adjustments was to attempt to account for changes over time in baseline characteristics of SARS-CoV-2 positive patients (resulting, for example, from increased availability of testing) that could have resulted in changes in the risk of adverse outcomes. Interrupted time series analysis of trends methods were used to test monthly trends in outcomes over the testing period from February through September 2020^36^. Time trends were modeled using ordinary least squares with adjustment for an auto-correlation lag of one month. Robust variances estimators were used to address heteroskedasticity.

## Results

### Trends in baseline characteristicsof patients with SARS-CoV-2 infection

Among all 55,952 individuals who tested positive for SARS-CoV-2 during the study period, the majority were male (89.3%), mean age was 60.0 years, 58.2% were White, 31.0% were Black, 12.3% were Hispanic, 12.4% had a CCI≥5, 25.4% had documentation of fever and 11% had documentation of dyspnea (**Table 1**).

**Table 1.** Baseline characteristics of VA patients who tested positive for SARS-CoV-2 between 2/28/20 and 9/30/20, by month

|  | All Patients<br>N=55,952 | Before March 31<br>N=2504 | April<br>N=7451 | May<br>N=5025 | June<br>N=7782 | July<br>N=16,273 | August<br>N=9284 | September<br>N=7633 |
| --- | --- | --- | --- | --- | --- | --- | --- | --- |
| **Sex** |  |  |  |  |  |  |  |  |
| Female (%) | 10.7 | 9 | 9.8 | 9.2 | 11.3 | 12.1 | 10.5 | 9.7 |
| Male (%) | 89.3 | 91 | 90.2 | 90.8 | 88.7 | 87.9 | 89.5 | 90.3 |
| **Age (years) mean (sd)** | 60.0±16.8 | 60.7±15.4 | 64.1±16.4 | 64.1±16.5 | 57.5±17.5 | 57.6±16.7 | 60.0±16.4 | 61.0±16.4 |
| 18-49 (%) | 27 | 24.2 | 19 | 18.3 | 33.1 | 32.4 | 26.2 | 25.1 |
| 50-64 (%) | 28.2 | 31.4 | 27.6 | 28.6 | 27.5 | 28.6 | 28.7 | 26.6 |
| 65-79 (%) | 34.1 | 35.1 | 37.2 | 36.5 | 30 | 31.2 | 35.5 | 37.6 |
| >=80 (%) | 10.7 | 9.3 | 16.1 | 16.7 | 9.5 | 7.8 | 9.6 | 10.7 |
| **Race** |  |  |  |  |  |  |  |  |
| White (%) | 58.2 | 39.2 | 51.9 | 55.8 | 58.1 | 57.9 | 61.9 | 68.2 |
| Black (%) | 31 | 51.9 | 39 | 34.3 | 29.9 | 30.3 | 27.5 | 21.3 |
| Other (%) | 2.8 | 2.2 | 2.3 | 2.7 | 3.2 | 3 | 2.9 | 2.9 |
| Missing/Unknown (%) | 8 | 6.7 | 6.9 | 7.2 | 8.8 | 8.8 | 7.7 | 7.6 |
| **Ethnicity** |  |  |  |  |  |  |  |  |
| Non-Hispanic (%) | 84.3 | 86.3 | 88.1 | 88.1 | 80.1 | 80.8 | 85.3 | 88.3 |
| Hispanic (%) | 12.3 | 11.1 | 8.8 | 8.6 | 16.1 | 15.8 | 11.1 | 8.5 |
| Missing/Unknown (%) | 3.4 | 2.6 | 3.2 | 3.3 | 3.8 | 3.5 | 3.5 | 3.2 |
| **Geographical Region\*** |  |  |  |  |  |  |  |  |
| 1 | 3.3 | 3.9 | 11.3 | 7.8 | 2.4 | 0.9 | 1.1 | 1.3 |
| 2 | 7 | 24.2 | 23 | 11.4 | 3.4 | 2 | 2.4 | 2.8 |
| 3 | 7.2 | 7.6 | 11.5 | 13.4 | 5.8 | 4.7 | 6.4 | 6.3 |
| 4 | 29.8 | 14.1 | 14.7 | 20.1 | 31.9 | 36.8 | 36.5 | 30.6 |
| 5 | 11.8 | 16.7 | 16.5 | 19.6 | 8.1 | 7.6 | 11 | 14.5 |
| 6 | 18.7 | 18.3 | 9.3 | 10 | 22.9 | 25.1 | 18.5 | 16.1 |
| 7 | 5.4 | 2.5 | 3.5 | 5 | 3.1 | 3.8 | 7.6 | 11.5 |
| 8 | 2.9 | 3.4 | 2.9 | 3.9 | 2.1 | 1.8 | 2.7 | 5.5 |
| 9 | 11.5 | 7.2 | 5.3 | 6.9 | 18.4 | 14.8 | 10.7 | 8.7 |
| 10 | 2.4 | 2.1 | 2 | 1.9 | 1.9 | 2.5 | 3.2 | 2.7 |
| **Urban vs. Rural** |  |  |  |  |  |  |  |  |
| Rural/Highly rural (%) | 36 | 18.8 | 21.4 | 28 | 32.6 | 36.9 | 45.1 | 51.9 |
| Urban (%) | 50.8 | 61.2 | 61 | 55.8 | 52.2 | 50 | 44.9 | 41.7 |
| Missing (%) | 13.1 | 19.9 | 17.6 | 16.2 | 15.2 | 13.1 | 10 | 6.4 |
| **BMI** |  |  |  |  |  |  |  |  |
| <18.5 (underweight) (%) | 1.2 | 1.5 | 1.9 | 2.1 | 1.2 | 0.7 | 1.1 | 0.9 |
| 18.5-24.9 (normal weight) (%) | 15.9 | 16.6 | 19.2 | 20.4 | 15.6 | 14.4 | 14.4 | 14.7 |
| 25-29.9 (overweight) (%) | 31.5 | 31.9 | 31.1 | 31.1 | 32.1 | 31.7 | 30.8 | 32 |
| 30-34.9 (Obese I) (%) | 27.6 | 27.1 | 25.2 | 24.9 | 27.8 | 28.5 | 28.9 | 28.5 |
| ≥35 (Obese II and III) (%) | 21.6 | 21.8 | 20.1 | 18.7 | 21.2 | 22.5 | 22.8 | 21.7 |
| Missing (%) | 2.2 | 1 | 2.5 | 2.8 | 2.2 | 2.2 | 2.1 | 2.2 |
| **Charlson Comorbidity Index (CCI)** |  |  |  |  |  |  |  |  |
| 0 (%) | 39.3 | 32.2 | 31.4 | 32.7 | 43.7 | 43.2 | 39.8 | 40.4 |
| 1-2 (%) | 33.6 | 34.2 | 32.4 | 33.7 | 32.7 | 34.1 | 34.4 | 33.5 |
| 3-4 (%) | 14.6 | 15.5 | 17.9 | 17 | 12.6 | 12.9 | 14.8 | 15.1 |
| ≥5 (%) | 12.4 | 18.1 | 18.3 | 16.6 | 11 | 9.7 | 11 | 11 |
| Fever (%) | 25.4 | 57.7 | 33.4 | 22.3 | 26.8 | 22.9 | 19.9 | 20 |
| Dyspnea(%) | 11 | 26.6 | 15.1 | 10.3 | 10.8 | 9.6 | 8.4 | 8.9 |
| Albumin (g/dL)† |  |  |  |  |  |  |  |  |
| > 3.9 | 24.6 | 29.1 | 19 | 18.7 | 28.1 | 25.7 | 26.2 | 25.7 |
| >3.6 to 3.9 | 19.4 | 19.7 | 16.8 | 18 | 18.4 | 20.6 | 20.1 | 21.9 |
| >3.1 to 3.6 | 30.4 | 30.2 | 31.9 | 27.9 | 30.9 | 31.2 | 29.5 | 29.3 |
| >2.7 to 3.1 | 15.2 | 13.6 | 18.2 | 20.1 | 13.6 | 13.2 | 14.8 | 14.4 |
| ≤ 2.7 | 10.4 | 7.5 | 14.2 | 15.4 | 9 | 9.3 | 9.4 | 8.6 |
| Aspartate aminotransferase† |  |  |  |  |  |  |  |  |
| ≤ 23 | 27 | 16.5 | 22.9 | 31.9 | 29.5 | 26.7 | 30.5 | 32.2 |
| >23 to 33 | 24.1 | 23.6 | 22.3 | 21.1 | 27.8 | 25.2 | 24.5 | 23 |
| >33 to 49 | 23.5 | 24.4 | 24.4 | 25.3 | 21.9 | 23.2 | 23.8 | 21.8 |
| >49 to 78 | 15.3 | 21.1 | 17 | 13.3 | 13.1 | 14.8 | 13.2 | 15.2 |
| > 78 | 10.1 | 14.4 | 13.4 | 8.4 | 7.8 | 10 | 8 | 7.8 |
| Creatinine (mg/dL) † |  |  |  |  |  |  |  |  |
| ≤ .95 | 25.3 | 20.3 | 23.4 | 28.3 | 27.3 | 25.4 | 26.3 | 26.9 |
| >.95 to 1.2 | 25.6 | 27.7 | 24.5 | 24.7 | 25 | 25.3 | 26.1 | 26.6 |
| >1.2 to 1.67 | 23.2 | 23.4 | 22.7 | 21 | 23 | 24.1 | 23.3 | 23.8 |
| >1.67 to 2.99 | 15.4 | 17.4 | 16.1 | 14.8 | 14.4 | 15.5 | 15.6 | 13.8 |
| > 2.99 | 10.5 | 11.3 | 13.3 | 11.3 | 10.3 | 9.7 | 8.7 | 8.9 |
| White blood cell count (k/μL)† |  |  |  |  |  |  |  |  |
| ≤ 4.7 | 25.3 | 30.1 | 23 | 24.1 | 26.2 | 26.5 | 23.5 | 24.1 |
| >4.7 to 6.2 | 25.3 | 27.8 | 24.4 | 26.2 | 24.2 | 26.6 | 24.2 | 23.8 |
| >6.2 to 8.32 | 24.5 | 23.7 | 25.3 | 24.4 | 25.3 | 23.2 | 24.6 | 25.5 |
| >8.32 to 11.3 | 15 | 10.9 | 17.2 | 14.8 | 15.6 | 14.4 | 15.6 | 15.5 |
| > 11.3 | 9.9 | 7.5 | 10.1 | 10.4 | 8.7 | 9.4 | 12.1 | 11.2 |
| Neutrophil-to-lymphocyte ratio (sd) † |  |  |  |  |  |  |  |  |
| ≤ 2.46 | 26 | 20.5 | 20.6 | 27.3 | 31.6 | 28.7 | 27.3 | 25.4 |
| >2.46 to 4.2 | 25 | 26.3 | 23.5 | 25.5 | 25.2 | 25.6 | 23.2 | 26.1 |
| >4.2 to 7.17 | 24.7 | 29.6 | 25.9 | 23.2 | 22 | 24.3 | 24.7 | 23.2 |
| >7.17 to 12.25 | 14.7 | 15.4 | 18.7 | 14.6 | 12.9 | 13.2 | 13.3 | 15.1 |
| > 12.25 | 9.6 | 8.3 | 11.3 | 9.4 | 8.2 | 8.2 | 11.5 | 10.2 |

\*Categorized according to the 10 “Standard Federal Regions” drawn up by the Office of Management and Budget: 1 (CT, MA, ME, NH, RI, VT), 2 (NJ, NY, PR), 3 (DC, DE, MD, PA, VA, WV), 4 (AL, FL, GA, KY, MS, NC, SC, TN), 5 (IL, IN, MI, MN, OH, WI), 6 (AR, LA, NM, OK, TX), 7 (IA, KS, MO, NE), 8 (CO, MT, ND, SD, UT, WY), 9 (AZ, CA, GU, HI, NV), 10 (AK, ID, OR, WA from https://www.fema.gov/about/organization/regions).
†Laboratory tests are reported for hospitalized patients only and categorized according to percentiles for all patients: 0-25, 25-50, 50-75, 75-90 and 90-100

The number of patients testing positive each month ranged from 13,967 in the earliest time period (2/28/20 to 03/31/20) to 124,771 in July. There were substantial trends in certain baseline socio-demographic characteristics of patients who tested positive for SARS-CoV-2 across successive months, including a decrease over time in the proportion who were Black (from 51.9% before March 31 to 21.3% in September), an increase in the proportion who were white (from 51.9% to 68.2%) and an increase in the proportion who resided in rural/highly rural (vs. urban) areas (from 18.8% to 51.9% in September). The proportion of patients originating from each U.S. geographical region varied dramatically over time consistent with well-described changes in disease epicenters (e.g 24.2% of positive patients from region 2 (NY, NJ, PR) in February/March versus only 2.8% in September).

The proportion of patients with severe comorbidity burden (CCI≥5) decreased over time (from 18.1% to 11%) as did the proportion with documented fever (from 57.7% to 20%) or dyspnea (from 26.6% to 8.9%). Among patients who were hospitalized, there were no clear temporal trends the proportion who had abnormalities in the five laboratory tests examined.

### Trends in pharmacotherapy, dialysis and non-invasive ventilation in patients with SARS-CoV-2 infection

The proportion of hospitalized patients who received hydroxychloroquine on or after the index date decreased from 56.6% in February/March to 2.4% in May 2020 and ranged from 0-0.6% thereafter (**Table 2, Figure 1a**). The proportion of hospitalized patients prescribed azithromycin decreased from 48.3% in February/March to 16.6% in September and use of vasopressors decreased from 20.6% to 8.7%. Tocilizumab prescription rates in hospitalized patients ranged from a peak of 8.7% in April to 0.8% in September 2020. In contrast, remdesivir prescriptions increased from 1.7% to 45.4% of hospitalized patients from February/March to September. Dexamethasone prescriptions were low during the first three months of the ascertainment period (<4% for hospitalized patients) but from May to September increased from 3% to 53.1% among hospitalized patients and prescription of corticosteroids other than dexamethasone also increased during the same time from 7.8% to 29%. Prescription of anticoagulants among hospitalized patients did not change appreciably over time. Use of investigational therapies including eculizumab and androgen deprivation therapy such as degarelix was minimal throughout the ascertainment period. The percentage of hospitalized patients who received dialysis decreased from a high of 11.6% in the earliest time period to a low of 3.8% in August, and use of non-invasive ventilation increased from a low of 8.4% in February/March to a high of 12.5% in August.

**Figure 1.**
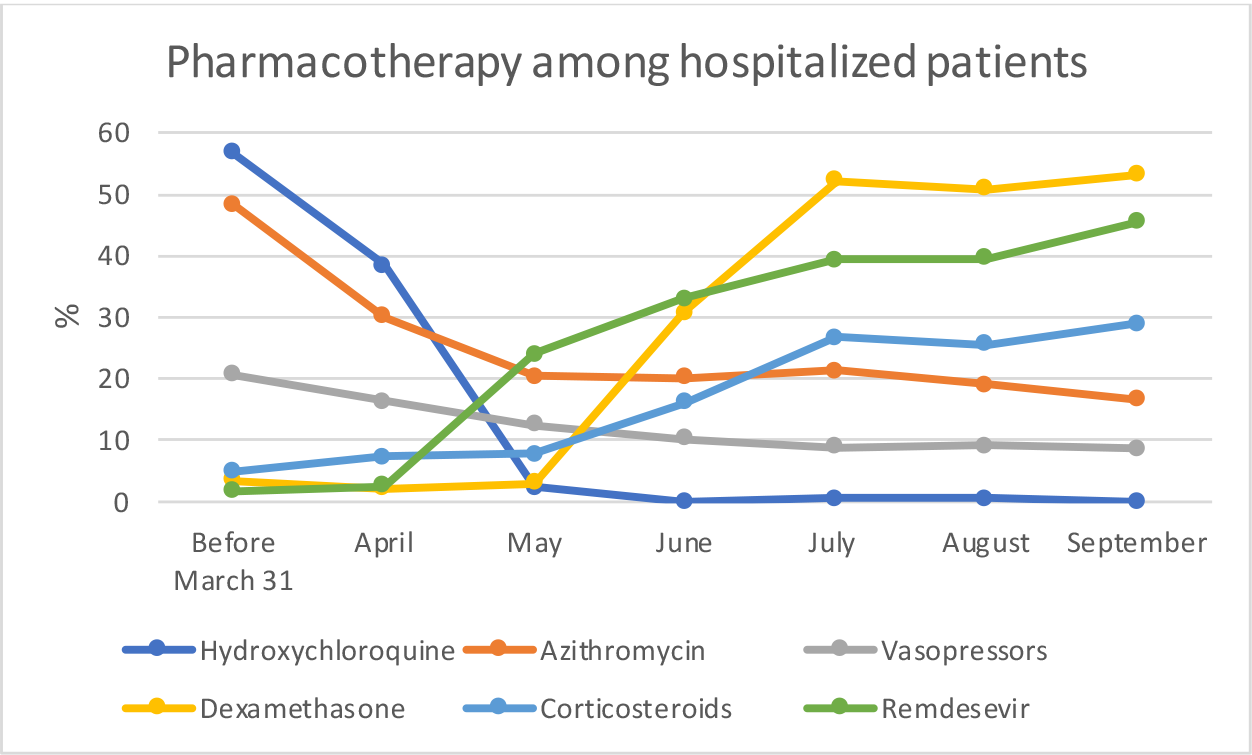
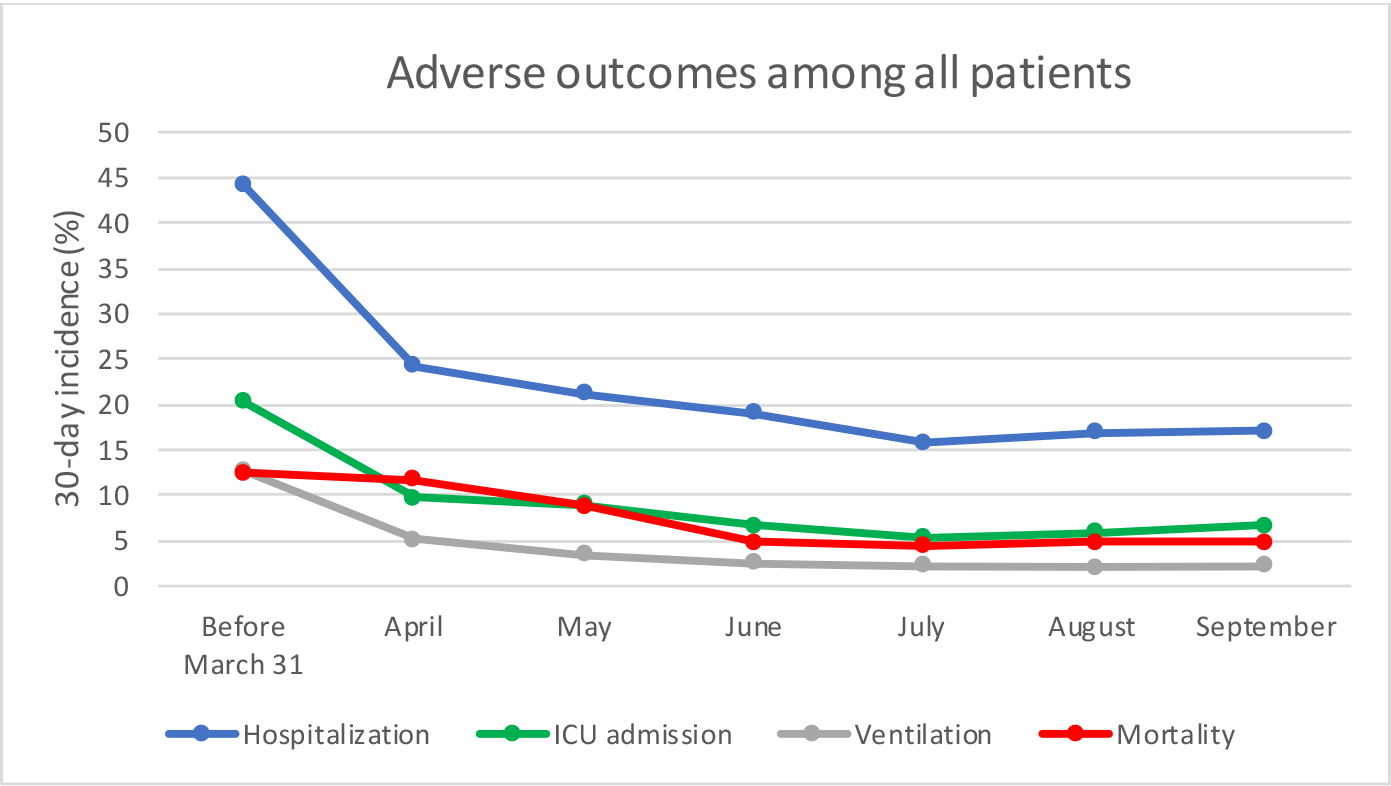
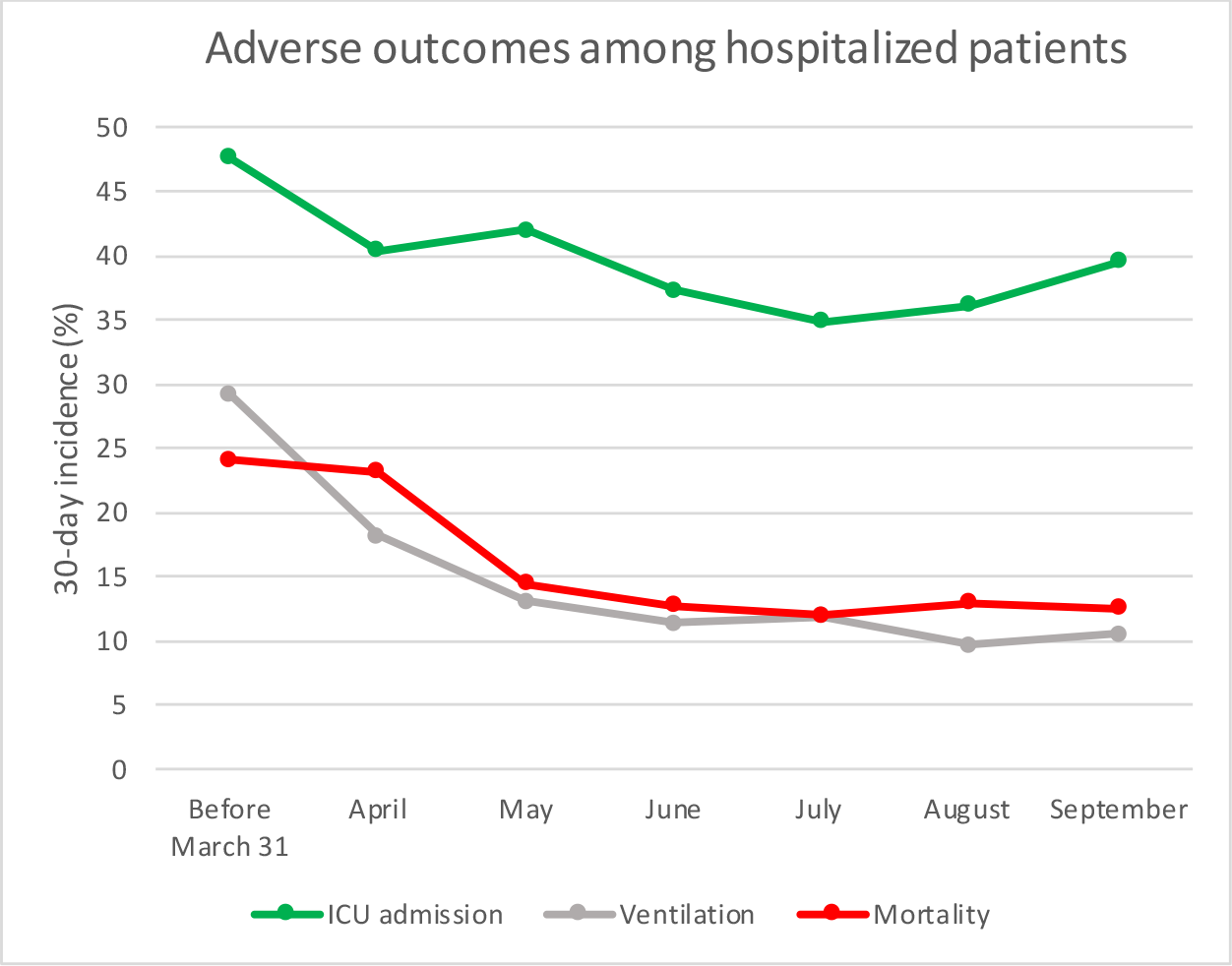
**Trends over time in pharmacotherapy and adverse outcomes of SARS-CoV-2 infection in a national cohort of VA patients** **1a. Trends over time in pharmacotherapy among SARS-CoV-2-infected hospitalized patients** (“Vasopressors” include Norepinephrine, Dobutamine, Vasopressin) **1b. Trends over time in 30-day adverse outcomes among all SARS-CoV-2-infected patients** **1c. Trends over time in 30-day adverse outcomes among SARS-CoV-2-infected hospitalized patients**

**Table 2.** Changes in pharmacotherapy, noninvasive ventilation and dialysis over time in patients with SARS-CoV-2 infection

|  | All Patients<br>N=55,952 | Before March<br>31<br>N=2504 | April<br>N=7451 | May<br>N=5025 | June<br>N=7782 | July<br>N=16,273 | August<br>N=9284 | September<br>N=7633 |
| --- | --- | --- | --- | --- | --- | --- | --- | --- |
|  | **ALL PATIENTS** |  |  |  |  |  |  |  |
| **MEDICATIONS\*** |  |  |  |  |  |  |  |  |
| **Decreasing prescriptions** |  |  |  |  |  |  |  |  |
| Hydroxychloroquine (%) | 2.6 | 26.2 | 9.5 | 0.5 | 0.1 | 0.2 | 0.2 | 0.1 |
| Azithromycin (%) | 7.9 | 26.5 | 9.5 | 5.9 | 7.2 | 6.9 | 6.5 | 5.9 |
| Tocilizumab (%) | 0.8 | 2.6 | 2.1 | 1.1 | 0.7 | 0.6 | 0.2 | 0.1 |
| Vasoressors‡ (%) | 2.8 | 9.3 | 4.4 | 3.2 | 2.4 | 1.8 | 1.9 | 1.9 |
| **Increasing prescriptions** |  |  |  |  |  |  |  |  |
| Remdesivir (%) | 5.4 | 0.7 | 0.7 | 4.9 | 6.3 | 6.2 | 6.5 | 7.6 |
| Dexamethasone (%) | 7.0 | 1.6 | 0.5 | 0.8 | 6.7 | 9.5 | 9.8 | 10.5 |
| Corticosteroids (%)†† | 5.8 | 4.6 | 3.4 | 2.8 | 5.4 | 6.9 | 6.7 | 7.7 |
| **Rare prescriptions** |  |  |  |  |  |  |  |  |
| Ecuzumab (%) | 0.0 | 0.0 | 0.0 | 0.0 | 0.0 | 0.0 | 0.0 | 0.0 |
| ADT (%) † | 0.0 | 0.0 | 0.1 | 0.0 | 0.0 | 0.1 | 0.0 | 0.0 |
| Degarelix (%) | 0.0 | 0.0 | 0.0 | 0.0 | 0.0 | 0.0 | 0.0 | 0.0 |
| **Unchanged prescriptions** |  |  |  |  |  |  |  |  |
| Anticoagulants (%) | 11.5 | 24.4 | 13.7 | 12.0 | 11.4 | 10.0 | 10.2 | 9.8 |
| **DIALYSIS\*\* (%)** | 1.5 | 5.2 | 2.3 | 1.9 | 1.2 | 1.2 | 0.9 | 1.0 |
| **NON-INVASIVE VENTILATION (%)** | 3.0 | 4.5 | 2.9 | 2.9 | 3.0 | 2.9 | 3.1 | 2.8 |
|  | **HOSPITALIZED PATIENTS** |  |  |  |  |  |  |  |
|  | N= 9,294 | N= 979 | N= 1,581 | N= 930 | N= 1,203 | N= 2,181 | N= 1,314 | N= 1,106 |
| **MEDICATIONS** |  |  |  |  |  |  |  |  |
| **Decreasing prescriptions** |  |  |  |  |  |  |  |  |
| Hydroxychloroquine (%) | 13.0 | 56.6 | 38.4 | 2.4 | 0.0 | 0.6 | 0.6 | 0.0 |
| Azithromycin (%) | 24.5 | 48.3 | 30.0 | 20.3 | 20.2 | 21.4 | 19.1 | 16.6 |
| Tocilizumab (%) | 4.4 | 5.6 | 8.7 | 5.1 | 4.0 | 4.4 | 1.1 | 0.8 |
| Vasoressors ‡ (%) | 11.9 | 20.6 | 16.4 | 12.4 | 10.3 | 8.9 | 9.1 | 8.7 |
| **Increasing prescriptions** |  |  |  |  |  |  |  |  |
| Remdesivir (%) | 27.5 | 1.7 | 2.5 | 24.0 | 33.0 | 39.3 | 39.6 | 45.4 |
| Dexamethasone (%) | 30.7 | 3.4 | 2.1 | 3.0 | 30.8 | 52.1 | 50.9 | 53.1 |
| Corticosteroids (%)†† | 17.9 | 4.9 | 7.3 | 7.8 | 16.0 | 26.6 | 25.6 | 29.0 |
| **Rare prescriptions** |  |  |  |  |  |  |  |  |
| Ecuzumab (%) | 0.0 | 0.0 | 0.0 | 0.0 | 0.0 | 0.0 | 0.0 | 0.0 |
| ADT (%)† | 0.2 | 0.0 | 0.1 | 0.1 | 0.0 | 0.3 | 0.3 | 0.2 |
| Degarelix (%) | 0.1 | 0.0 | 0.0 | 0.0 | 0.0 | 0.1 | 0.2 | 0.1 |
| **Unchanged prescriptions** |  |  |  |  |  |  |  |  |
| Anticoagulants (%) | 51.8 | 52.5 | 50.0 | 49.7 | 51.6 | 54.8 | 51.5 | 50.3 |
| **DIALYSIS (%)** | 6.0 | 11.6 | 7.7 | 5.7 | 4.6 | 5.3 | 3.8 | 4.2 |
| **NON-INVASIVE VENTILATION (%)** | 10.7 | 8.4 | 9.4 | 9.5 | 10.6 | 12.2 | 12.5 | 11.2 |

\*Medications refer to new prescriptions of medications on or within 60 days after the index date that were not prescribed before the index date.
†ADT = Androgen Deprivation Therapy
‡ Vasoressors: Norepinephrine, Dobutamine, Vasopressin
\*\* Dialysis refers to new initiation of dialysis on or after the index date in patients who were not on dialysis before
†† Corticosteroids, other than dexamethasone

Analogous decreases in prescription of hydroxychloroquine, azithromycin and vasopressors and increases in prescription of remdesivir, dexamethasone and other corticosteroids were observed when presented as a proportion of all infected patients (rather than the hospitalized subgroup) – **Table 2**.

### Trends in risk of adverse outcomes: hospitalization, ICU admission, mechanical ventilation, death

Between February and July 2020, there were downward trends in the 30-day incidence of hospitalization (44.2% to 15.8%), ICU admission (20.3% to 5.3%), mechanical ventilation (12.7% to 2.2%), and death (12.5% to 4.4%) among patients who tested positive for SARS-CoV-2, with relative stabilization from July through September, 2020 (15.8% to 17.1% for hospitalization; 5.3% to 6.6% for ICU admission; 2.2% to 2.2% for mechanical ventilation; and 4.4% to 4.9% for mortality) (**Table 3, Figure 1b**).

**Table 3.** Association between time period and adverse outcomes in patients with SARS-CoV-2 infection

| Dates of SARS-CoV-2 testing | Number positive<br>N=55,952 | Number with Event | 30-day Incidence | Unadjusted Hazard Ratio | Adjusted Hazard Ratio* | p-value for trends**/† |
| --- | --- | --- | --- | --- | --- | --- |
| **ALL PATIENTS** |  |  |  |  |  |  |
|  |  | **Hospitalization** |  |  |  |  |
| Before March 31 | 2,504 | 1,104 | 44.2 | 1 | 1 | < 0.001/0.04 |
| April | 7,451 | 1,798 | 24.3 | 0.53(0.49-0.57) | 0.66(0.61-0.71) |  |
| May | 5,025 | 1,057 | 21.2 | 0.47(0.43-0.51) | 0.68(0.62-0.74) |  |
| June | 7,782 | 1,472 | 19.0 | 0.40(0.37-0.44) | 0.68(0.63-0.75) |  |
| July | 16,273 | 2,567 | 15.8 | 0.35(0.32-0.37) | 0.62(0.57-0.67) |  |
| August | 9,284 | 1,561 | 16.9 | 0.37(0.34-0.40) | 0.64(0.59-0.70) |  |
| September | 7,633 | 1,305 | 17.1 | 0.38(0.35-0.41) | 0.66(0.60-0.72) |  |
|  |  | **ICU admission** |  |  |  |  |
| Before March 31 | 2,504 | 506 | 20.3 | 1 | 1 | < 0.001/0.03 |
| April | 7,451 | 708 | 9.7 | 0.45(0.40-0.50) | 0.56(0.49-0.63) |  |
| May | 5,025 | 446 | 9.0 | 0.41(0.36-0.47) | 0.61(0.53-0.69) |  |
| June | 7,782 | 521 | 6.7 | 0.33(0.29-0.37) | 0.56(0.49-0.64) |  |
| July | 16,273 | 863 | 5.3 | 0.27(0.24-0.31) | 0.49(0.43-0.55) |  |
| August | 9,284 | 548 | 5.9 | 0.28(0.25-0.32) | 0.50(0.44-0.57) |  |
| September | 7,633 | 499 | 6.6 | 0.32(0.28-0.36) | 0.56(0.49-0.65) |  |
|  |  | **Mechanical Ventilation** |  |  |  |  |
| Before March 31 | 2,504 | 316 | 12.7 | 1 | 1 | < 0.001/0.04 |
| April | 7,451 | 376 | 5.2 | 0.38(0.33-0.45) | 0.47(0.40-0.55) |  |
| May | 5,025 | 168 | 3.4 | 0.24(0.20-0.30) | 0.36(0.29-0.44) |  |
| June | 7,782 | 189 | 2.5 | 0.16(0.13-0.19) | 0.26(0.21-0.32) |  |
| July | 16,273 | 351 | 2.2 | 0.15(0.12-0.17) | 0.25(0.21-0.29) |  |
| August | 9,284 | 191 | 2.1 | 0.14(0.11-0.17) | 0.23(0.19-0.28) |  |
| September | 7,633 | 164 | 2.2 | 0.15(0.12-0.18) | 0.25(0.20-0.31) |  |
|  |  | **Mortality** |  |  |  |  |
| Before March 31 | 2,504 | 312 | 12.5 | 1 | 1 | < 0.001/< 0.01 |
| April | 7,451 | 874 | 11.7 | 0.84(0.73-0.96) | 0.73(0.63-0.83) |  |
| May | 5,025 | 442 | 8.8 | 0.62(0.53-0.72) | 0.52(0.45-0.61) |  |
| June | 7,782 | 378 | 4.9 | 0.35(0.30-0.41) | 0.42(0.35-0.49) |  |
| July | 16,273 | 719 | 4.4 | 0.32(0.28-0.37) | 0.40(0.34-0.46) |  |
| August | 9,284 | 441 | 4.8 | 0.34(0.29-0.39) | 0.37(0.32-0.44) |  |
| September | 7,633 | 373 | 4.9 | 0.34(0.29-0.41) | 0.38(0.32-0.45) |  |
| **HOSPITALIZED PATIENTS** |  |  |  |  |  |  |
|  |  | **ICU admission** |  |  |  |  |
| Before March 31 | 979 | 462 | 47.6 | 1 | 1 | < 0.001/< 0.01 |
| April | 1,581 | 631 | 40.4 | 0.80(0.71-0.91) | 0.75(0.66-0.85) |  |
| May | 930 | 388 | 42.0 | 0.81(0.70-0.93) | 0.85(0.73-0.98) |  |
| June | 1,203 | 447 | 37.3 | 0.75(0.65-0.86) | 0.81(0.70-0.94) |  |
| July | 2,181 | 755 | 34.8 | 0.70(0.62-0.80) | 0.74(0.64-0.84) |  |
| August | 1,314 | 472 | 36.1 | 0.72(0.62-0.83) | 0.73(0.63-0.84) |  |
| September | 1,106 | 436 | 39.5 | 0.77(0.67-0.89) | 0.82(0.70-0.95) |  |
| October |  |  |  |  |  |  |
| November |  |  |  |  |  |  |
|  |  | **Mechanical Ventilation** |  |  |  |  |
| Before March 31 | 979 | 283 | 29.2 | 1 | 1 | < 0.001/0.04 |
| April | 1,581 | 282 | 18.2 | 0.60(0.50-0.71) | 0.56(0.47-0.66) |  |
| May | 930 | 120 | 13.1 | 0.39(0.31-0.49) | 0.45(0.36-0.57) |  |
| June | 1,203 | 135 | 11.4 | 0.30(0.24-0.38) | 0.35(0.27-0.44) |  |
| July | 2,181 | 255 | 11.9 | 0.31(0.26-0.38) | 0.33(0.27-0.40) |  |
| August | 1,314 | 125 | 9.7 | 0.26(0.20-0.33) | 0.26(0.21-0.33) |  |
| September | 1,106 | 114 | 10.5 | 0.29(0.23-0.36) | 0.32(0.25-0.41) |  |
|  |  | **Mortality** |  |  |  |  |
| Before March 31 | 979 | 236 | 24.1 | 1 | 1 | < 0.001/< 0.01 |
| April | 1,581 | 367 | 23.2 | 0.93(0.79-1.11) | 0.65(0.55-0.78) |  |
| May | 930 | 134 | 14.4 | 0.56(0.45-0.70) | 0.44(0.35-0.55) |  |
| June | 1,203 | 153 | 12.7 | 0.49(0.39-0.61) | 0.45(0.35-0.57) |  |
| July | 2,181 | 261 | 12.0 | 0.45(0.37-0.55) | 0.39(0.31-0.48) |  |
| August | 1,314 | 170 | 12.9 | 0.49(0.39-0.61) | 0.38(0.30-0.48) |  |
| September | 1,106 | 138 | 12.5 | 0.47(0.37-0.59) | 0.38(0.30-0.49) |  |
| **ICU ADMITTED PATIENTS** |  |  |  |  |  |  |
|  |  | **Mechanical Ventilation** |  |  |  | < 0.001/0.03 |
| Before March 31 | 429 | 242 | 57.0 | 1 | 1 |  |
| April | 560 | 240 | 43.7 | 0.81(0.67-0.98) | 0.79(0.65-0.97) |  |
| May | 339 | 91 | 27.4 | 0.50(0.39-0.65) | 0.56(0.42-0.73) |  |
| June | 406 | 100 | 25.2 | 0.37(0.28-0.48) | 0.40(0.30-0.53) |  |
| July | 637 | 172 | 27.5 | 0.40(0.32-0.51) | 0.40(0.31-0.50) |  |
| August | 421 | 91 | 22.0 | 0.34(0.26-0.45) | 0.32(0.24-0.43) |  |
| September | 396 | 93 | 23.9 | 0.36(0.28-0.47) | 0.38(0.29-0.51) |  |
| October |  |  |  |  |  |  |
| November |  |  |  |  |  |  |
|  |  | **Mortality** |  |  |  | < 0.001/0.01 |
| Before March 31 | 429 | 168 | 39.2 | 1 | 1 |  |
| April | 560 | 201 | 35.9 | 1.04(0.84-1.29) | 0.71(0.56-0.89) |  |
| May | 339 | 77 | 22.7 | 0.65(0.49-0.88) | 0.51(0.37-0.69) |  |
| June | 406 | 94 | 23.2 | 0.64(0.48-0.86) | 0.54(0.40-0.73) |  |
| July | 637 | 135 | 21.2 | 0.53(0.41-0.69) | 0.48(0.36-0.63) |  |
| August | 421 | 95 | 22.6 | 0.59(0.44-0.79) | 0.44(0.32-0.60) |  |
| September | 396 | 80 | 20.2 | 0.54(0.40-0.74) | 0.46(0.33-0.63) |  |
| **PATIENTS WHO WERE MECHANICALLY VENTILATED** |  |  |  |  |  |  |
|  |  | **Mortality** |  |  |  | < 0.001/0.07 |
| Before March 31 | 247 | 144 | 58.3 | 1 | 1 |  |
| April | 245 | 138 | 56.3 | 1.04(0.80-1.36) | 0.85(0.64-1.11) |  |
| May | 96 | 30 | 31.3 | 0.57(0.37-0.88) | 0.48(0.30-0.76) |  |
| June | 108 | 43 | 39.8 | 0.80(0.53-1.21) | 0.63(0.41-0.98) |  |
| July | 180 | 79 | 43.9 | 0.98(0.68-1.40) | 0.86(0.59-1.27) |  |
| August | 90 | 42 | 46.7 | 0.99(0.63-1.53) | 0.81(0.51-1.29) |  |
| September | 78 | 33 | 42.3 | 0.77(0.49-1.22) | 0.63(0.39-1.02) |  |

\* FOR **ALL PATIENTS**: Adjusted for age, sex, race (White, Black, other, or missing/unknown), ethnicity, BMI, regional COVID-19 disease burden, Charlson comorbidity index, diabetes, chronic kidney disease, hypertension, obstructive sleep apnea, obesity hypoventilation, fever, and dyspnea (stratified by VA facility)
\* FOR **HOSPITALIZED, ICU ADMITTED AND VENTILATED PATIENTS** : Adjusted for age, sex, race, ethnicity, BMI, regional COVID-19 disease burden, Charlson comorbidity index, diabetes, chronic kidney disease, hypertension, obstructive sleep apnea, obesity hypoventilation, fever, dyspnea and the following laboratory tests: albumin, aspartate aminotransferase, creatinine, white blood cell count, neutrophil-to-lymphocyte ratio (stratified by VA facility).
\*\*Non-parametric Wilcoxon-type test for trend across ordered groups
†Interrupted time series test for difference in 30-day outcome rate slope before and after July, 2020.

Among hospitalized patients, the crude 30-day incidence of ICU admission decreased from 47.6% in February/March to 34.8% in July then increased slightly between July and September (34.8% to 39.5%) (**Table 3, Figure 1c**). The 30-day incidence of mechanical ventilation decreased from 29.2% in February/March to 11.4% in June and ranged between 9.7 and 11.9% from July to September. The 30-day incidence of mortality decreased from 24.1% in February/March to 12.0% in July and ranged between 12.5% to 12.9% thereafter. Among those admitted to the ICU, the crude 30-day incidence of mechanical ventilation ranged from 57.0% in February/March to 22.0% in August, increasing slightly to 23.9% in September. The crude 30-day mortality among those admitted to the ICU decreased from 39.2% in February/March to 22.7% in May, ranging from 20.2% to 23.2% thereafter. Among patients who received mechanical ventilation, the crude 30-day incidence of death decreased from 58.3% in February/March to 31.3% in May, increasing thereafter to between 39.8% to 46.7%. After adjustment for sociodemographic characteristics, comorbid conditions, symptoms and baseline laboratory characteristics, risk for all adverse outcomes decreased most markedly during the early months of the pandemic with stabilization during late summer and early fall (p<0.05 for interrupted time series test for difference in 30-day outcome rate slope before and after July, 2020 for all outcomes except for mortality among mechanically ventilated patients) (**Table 3 and Figure 2**).

**Figure 2.**
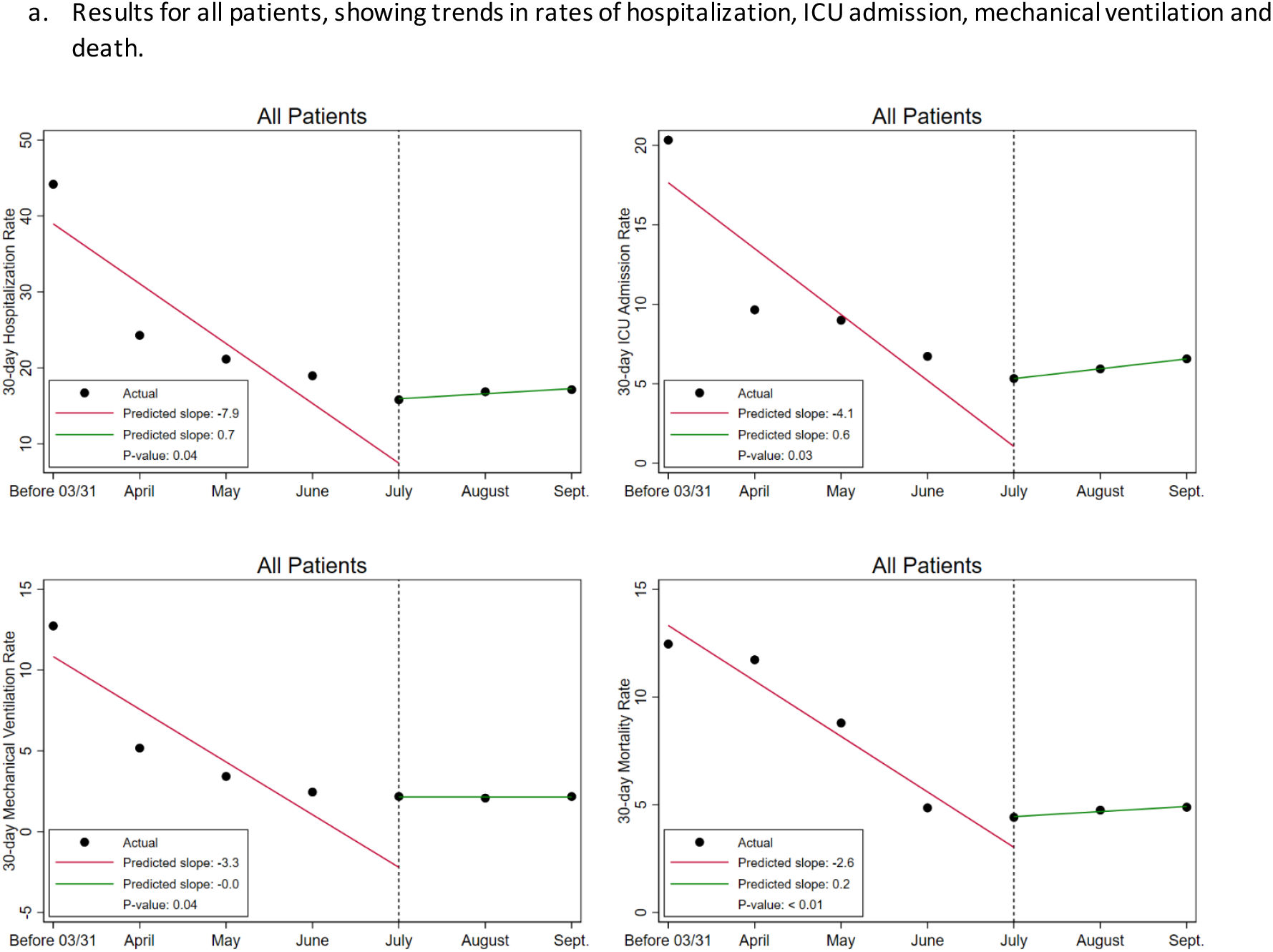
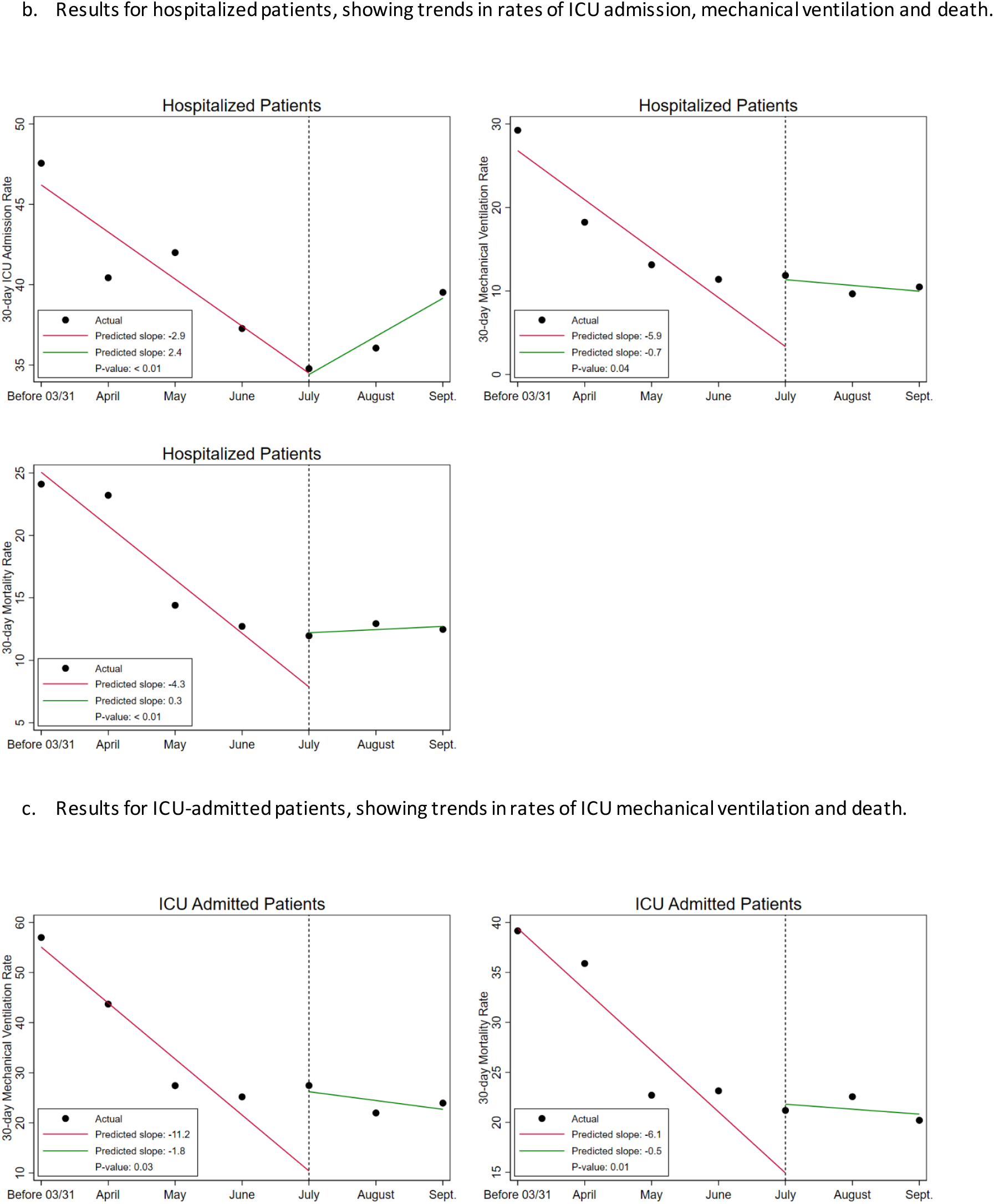
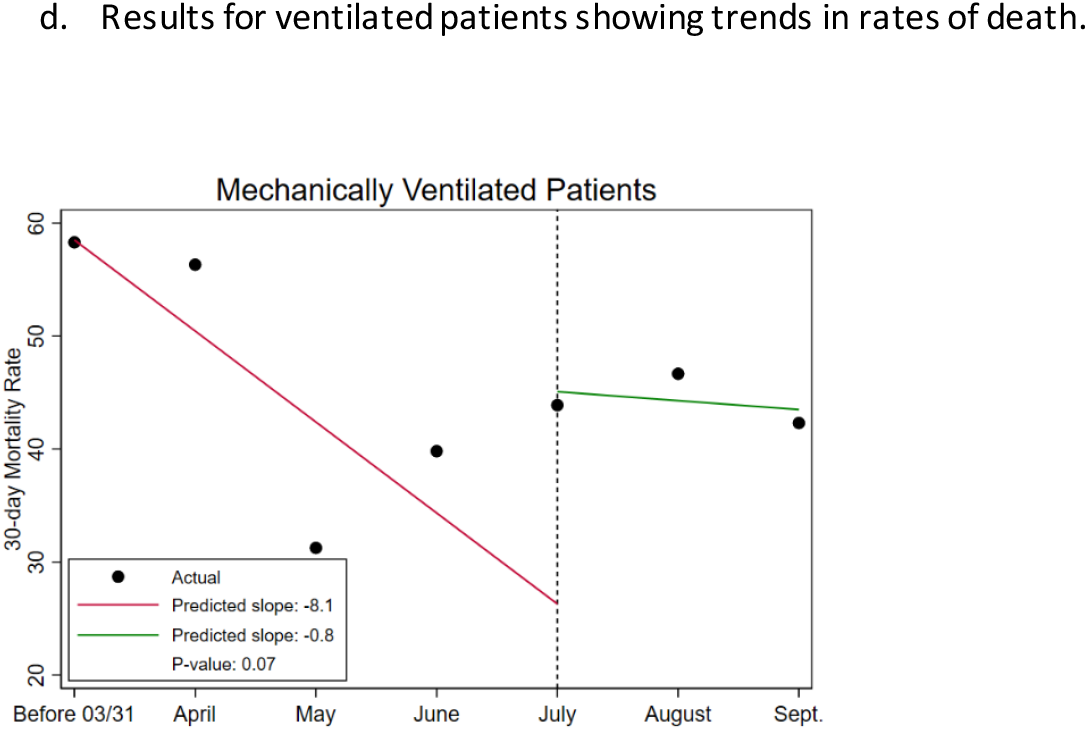
Graphical results of interrupted time series analysis for difference in 30-day outcome rate slope before and after July, 2020.

## Discussion

In our study of 55,952 VA enrollees who tested positive for SARS-CoV-2 between February 28 and September 30, 2020, there were marked decreases in the 30-day incidence of hospitalization, ICU admission, mechanical ventilation and death during the first wave of the US pandemic from February/March through July, 2020 with stabilization during the late summer and early fall. Similar trends were noted in analyses adjusted for baseline sociodemographic characteristics, comorbid conditions, symptoms and laboratory tests (among hospitalized patients) and in all subgroups examined, including those admitted to the hospital, those admitted to the ICU and those who received mechanical ventilation

While there have been marked changes over time in the characteristics of Veterans infected with SARS-CoV-2 that could perhaps have explained our findings, trends in improved outcomes during the first wave of the US pandemic with subsequent stabilization persisted after adjustment for a wide range of measured sociodemographic characteristics and comorbid conditions. While increased availability of testing over time would be expected to identify a greater number of asymptomatic or mildly symptomatic individuals infected with SARS-CoV-2, improvements in adverse outcomes during the initial months of the pandemic persisted after stratification by illness severity and among the sickest cohort members (i.e., those who were hospitalized, admitted to the ICU or received mechanical ventilation). It is also noteworthy that among hospitalized patients, results were similar after adjustment for a range of laboratory markers for illness acuity.

While it is possible that temporal trends in the frequency of adverse outcomes over time might reflect geographic variation in unmeasured factors (e.g., treatment practices) as the pandemic moved across the country, multivariable models were stratified by VA facility and our results were robust to adjustment for geographic region. Another possibility is that our results reflect changing thresholds for hospitalization, ICU admission and mechanical ventilation over the course of the pandemic. While relaxation in criteria for hospitalization, ICU admission and mechanical ventilation over the pandemic would be expected to be associated with reductions in mortality among patients who were hospitalized, admitted to the ICU or mechanically ventilated, we saw a decrease rather than increase in rates of each of these outcomes over time.

Our findings do raise the question of whether marked improvements in outcomes among patients infected with SARS-CoV-2 during the first wave of the pandemic might reflect changes in the clinical approach to caring for members of this population.^37^ We did observe very substantial increases in the use of potentially effective pharmacotherapy (e.g. remdesivir and dexamethasone) and decreases in the use of ineffective (e.g. azithromycin) or potentially harmful (e.g. hydroxychloroquine) therapy toward the end of the first wave of the pandemic, although inflection points in trends in the use of each of these agents did not exactly parallel those for adverse outcomes. Perhaps most importantly, there were probably other shifts in clinical practice that occurred within the first months of the pandemic that could not be reliably captured in the data sources available to us. Such shifts include high flow nasal canula [HFNC], pronation and noninvasive positive-pressure ventilation (NIPPV) which were increasingly used to avoid or delay intubation and may have resulted in improved outcomes^38^.

Another possible explanation for declining rates of adverse outcomes over time is that the severity of disease caused by SARS-CoV-2 infection may have declined between February and June 2020. This might occur, for example, through reduction in inoculum related to physical distancing or wearing face masks, which became more common during that time period.^39^ Also, as an RNA virus, SARS-CoV-2 does undergo rapid mutation and these mutations could impact the pathogenicity and clinical characteristics of infection^40, 41^. It is now clear that a SARS-CoV-2 variant carrying the Spike protein amino acid change D614G became progressively more common after March 2020 throughout the world, including the United States, and was the most prevalent variant by July 2020^42^. This strain has enhanced replication and is more infectious^42, 43^ with but may be associated with decreased disease severity. It is important to emphasize that such a reduction in disease severity caused by SARS-CoV-2 has not been proven, but it could explain declining trends in adverse outcomes over time that persisted after adjustment for baseline characteristics. Since the end of our study period, many viral variants have been described, such as the “United Kingdom variant” (B.1.1.7), the “South Africa variant” (B.1.351) and the “Brazil variant” (P.1), all of which have now been detected in the US. All of these variants have greater transmissibility, but it is not clear whether they are associated with worse disease severity or higher risk of death.

There are several limitations to our study. First, while use of a Veteran cohort provided a useful window on trends in complications of SARS-CoV-2 over time within a racially diverse national health care system, our cohort includes relatively few women which may limit the generalizability of our findings. We did not capture episodes of hospitalization, ICU admission or mechanical ventilation of VA patients in non-VA facilities (unless they were paid for by the VA), but there is no evidence that such non-VA care increased substantially over the course of our study. There may also have been a delay in capturing episodes of care occurring in the community under VA purchased care. However, this would not explain the temporal trends in adverse outcomes among hospitalized patients reported here. Finally, although we adjusted for a number of sociodemographic characteristics, comorbid conditions, symptoms and laboratory tests, some residual confounding by disease severity and other unmeasured factors likely persisted.

In conclusion, we identified marked improvements in outcomes of SARS-CoV-2 infection among Veterans infected during the first wave of the US pandemic with stabilization in late summer and early fall. These trends may reflect changing treatment practices, public health measures and viral pathogenicity. Studies should continue to track trends in adverse outcomes of SARS-CoV-2 infection.

## Data Availability

Data are not available due to restrictions imposed by the Department of Veterans Affairs on access to VA data.

## Acknowledgements

We acknowledge the VA Informatics and Computing Infrastructure (VINCI) group who worked tirelessly to create the “COVID 19 Shared Data Resource”.

